# Longitudinal viscosity of blood plasma for rapid COVID-19 prognostics

**DOI:** 10.1101/2023.10.13.23297016

**Authors:** Jennifer Illibauer, Tamara Clodi-Seitz, Alexander Zoufaly, Judith H. Aberle, Wolfgang J. Weninger, Manuela Foedinger, Kareem Elsayad

## Abstract

Blood Plasma Viscosity (PV) is an established biomarker for numerous diseases. While PV colloquially refers to the *shear viscosity*, there is a second viscosity component--the *bulk viscosity*--that describes the irreversible fluid compressibility on short time scales. The bulk viscosity is acutely sensitive to solid-like suspensions, and obtainable via the longitudinal viscosity from acoustic attenuation measurements. Whether it has diagnostic value remains unexplored yet may be pertinent given the association of diverse pathologies with the formation of plasma suspensions, such as fibrin-microstructures in COVID-19 and long-COVID. Here we show that the longitudinal PV measured using Brillouin Light Scattering (BLS) can serve as a proxy for the shear PV of blood plasma, and exhibits a temperature dependence consistent with increased suspension concentrations in severe COVID-patient plasma. Our results open a new avenue for PV diagnostics based on the longitudinal PV, and show that BLS can provide a means for its clinical implementation.

---

A major component of blood (∼55% by volume) is blood plasma, a semi-transparent dense electrolyte and proteins solution. Changes in the viscosity of plasma are known indicators of numerous pathologies, however are less frequently used owing in part to large patient variability^1^. Plasma can usually be assumed to be a Newtonian fluid (have a shear-rate independent viscosity) however, studies have revealed elastic properties with viscous relaxation times on the order of milliseconds, attributed to constituent proteins^2^. While these relaxation times are too fast to affect fluid dynamics under most physiological conditions, they show that viscoelastic properties at short time scales are sensitive to the soluble and insoluble constituents, and thus may provide a means for assessing their abundance and physical properties. While it is challenging to probe viscoelastic properties at such short time scales using conventional viscometers and shear-plate rheometers^3,4^, one approach that can probe at very short time scales, which has been receiving renewed interest for biomedical applications, is Brillouin Light Scattering (BLS) spectroscopy^5-7^.

## BLS Spectroscopy

BLS spectroscopy measures the hypersonic velocity and attenuation in a label-free all-optical fashion via the inelastic scattering of a single-frequency probing laser from collective molecular vibrations^5-7^. The measured spectrum in a BLS experiment consists of an elastic scattering peak straddled by two Brillouin scattering peaks (centered at frequencies + *v*_*B*_ and −*v*_*B*_ relative to the elastic peak) that result from the creation and annihilation of longitudinal acoustic phonons^5^ (Fig 1A). From the fitted peak positions *v*_*B*_ and linewidth of these peaks *𝛤*_*B*_, parameters such as the longitudinal elastic modulus, longitudinal acoustic attenuation and longitudinal viscosity can be calculated (Methods). Two important aspects differentiate BLS derived viscoelastic properties from those obtained using conventional viscometers and rheometers.

Firstly, in fluids BLS typically probes the *longitudinal* viscosity^5^, which depends not only on the shear but also the compressive properties, and is given by:

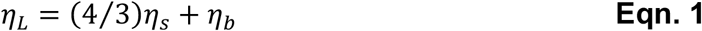

Here *η*_*s*_ is the shear viscosity and *η*_*b*_ the *bulk* (or *second*) viscosity^8^. Despite usually being about the same magnitude for fluids as *η*_*s*_, *η*_*b*_ is a distinct material property dependent on the constituent chemistry, such as the abundance and hydrostatic properties of solutes and suspensions^7^. *η*_*L*_ can also be measured using Acoustic Spectroscopy (AS) at lower (∼MHz) frequencies^9^, that unlike BLS, requires significantly larger sample volumes^9^ (>15mL compared to <100μL for BLS), which make it challenging for routine analysis of scarcely available biofluids. See Supplementary Table 1 for a list of differences between BLS and AS.

**Fig 1.**
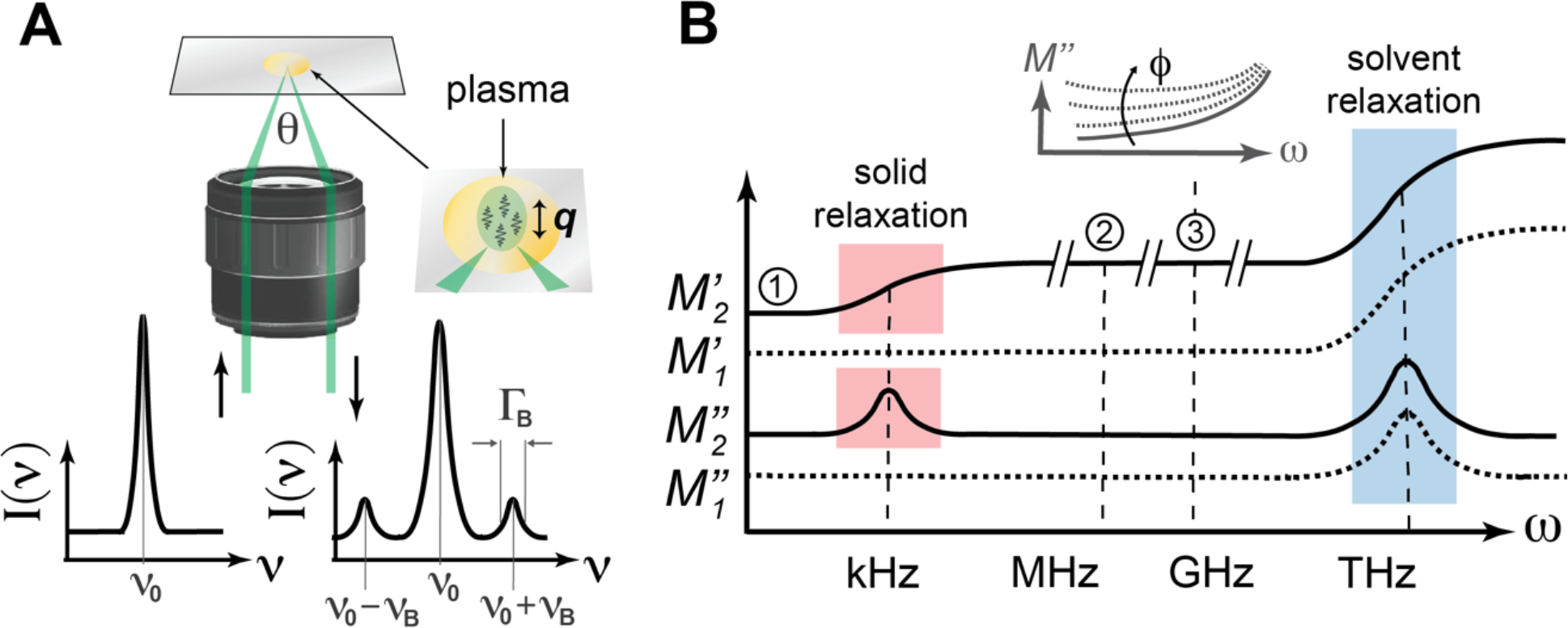
Brillouin light scattering (BLS) in liquids. **(A)** BLS spectroscopy measurement geometry and spectrum from an incident laser with frequency *v* = *v*_*B*_. The BLS scattering peaks are at *v* = *v*_*B*_ ± *v*_*L*_, with linewidth (Γ_*L*_). Probed phonon wavevector = ***q*. (B)** Sketch of the elastic and viscous longitudinal moduli frequency dispersion in a pure liquid 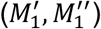 and a liquid with a solid-like suspension 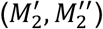. ➀, ➁ and ➂ indicate the frequencies probed using quasi-static techniques, acoustic spectroscopy, and BLS. *Inset:* expected change of the frequencydependence of *M*^″^ at ∼MHz-GHz frequencies for increasing suspension concentration (*ϕ*).

Secondly, BLS measures the viscoelastic properties at picosecond-nanosecond time scales—*i*.*e*. at the GHz frequencies of the probed phonons. To appreciate how this can differ from measurements at lower frequencies in liquids, Fig 1B shows a schematic of the frequency-dependence of the complex longitudinal modulus (*M* = *M*^′^ + *M*^″^*i*) in a *pure* fluid and a fluid with a solid-like suspension (*e*.*g*. proteins or aggregates thereof). Here *M*^′^ and *M*^″^ are the elastic (*storage*) and viscous (*loss*) contributions respectively. In both cases BLS probes the lower asymptotic limit of fast (>THz) solvent relaxation processes. However, while in a pure fluid, the BLS measured *M* is comparable to that at lower frequencies^9^, this is not the case if there is a suspension that contributes a slower relaxation process(es) 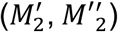. In addition to increasing *M*^′^, slower relaxation processes can also provide a contribution to the attenuation that counters the increasing contribution from the fast solvent relaxation (inset Fig 1B). Such an effect is also seen in ultrasonic measurements where the attenuation coefficient of blood plasma scales with frequency as:^10^ ∼ ω^1.6^, notably slower than the ω^2^ of pure liquids. While the dispersion depicted in Fig 1B serves as an approximation for a suspension solution, for blood plasma it can be expected to be significantly more complex with a spectrum of relaxation times dependent on those of its many constituents and their interactions.

## COVID and PV

Already early on in the COVID pandemic it was reported that acute COVID-19 infection is often associated with an increase in shear PV^11^. This was largely attributed to elevated fibrinogen levels in response to infection^11,12^. It has since become apparent that while COVID-19 infection manifests itself as a respiratory disease, its lethality usually stems from the disruption of coagulation processes^13^. Indeed, almost all COVID-19 critically ill ICU patients suffer from some form of clotting/lysis anomaly as diagnosed by Fibrin Degradation Product (FDP) blood values^14^. This insight—addressing it as a vascular/endothelial disease and coagulopathy^15,16^—shifted treatment strategies in ICUs around the world saving many lives.

Despite the clear markers of a coagulopathy, the puzzling thing that remained was that pathological analysis in many cases did not reveal the gross clots expected from blood chemistry (D-dimer) analysis^17^. It has been suggested that this is a result of the macrophage attack^18^, and gradual infusion of thrombin during infection into the pulmonary circulation^19^, resulting in soluble fibrin monomer complexes (SF) in the bloodstream (and thereby elevated FDP values). High levels of circulating SF may then lead to the formation of insoluble fibrin protofibrils and subsequently *microclots*^19^, which have been observed in COVID, as well as Long-COVID, patients^20^. The mobility of such microclots, their ability to cause transient blockage in narrow vasculature, together with a delayed fibrinolytic shutdown^21^, could then explain the multiple organ failure observed in severe COVID-19 patients^22^. A means for rapidly assessing changes in the physical properties of blood due to soluble or insoluble protein constituents, would thus be highly desirable for deciding and developing optimal treatment strategies and monitoring their effectiveness.

Considering the above, we here investigate whether the longitudinal PV measured using BLS can detect differences in the plasma between patients with different disease outcomes, how these scale with respect to the shear properties, and whether these can provide additional information indicative of disease progression.

## Results

Blood plasma from 39 hospitalized patients with PCR-confirmed SARS-CoV-2 infection and 20 PCR-confirmed SARS-CoV-2-negative and otherwise healthy persons was obtained and prepared as described in Methods. The COVID-positive samples were classed based on the World Health Organization (WHO) clinical progression scale, which is a measure of the implemented medical resources and disease trajectory^23^ (Methods). Given that PV is sensitive to a multitude of pathologies^1^ and lifestyle choices^24^ we focus on a subset of samples from non-smokers with none of the reported underlying conditions (Methods, Supplementary Table 2), as it is unclear how these would perturb results. For these, temperature sweep BLS and shear viscosity measurements were performed on a custom built BLS microscope and a commercial rolling-ball viscometer respectively, installed directly in a hospital blood bank (Methods). For BLS measurements, the fitted frequency-shift and linewidth after deconvolution, together with the separately measured refractive index and estimated mass density, were used to calculate the shear and longitudinal viscoelastic parameters (Methods). We found no correlation between viscoelastic parameters and patient age or gender (Supplementary Figure 1), consistent with previous shear viscosity studies on healthy plasma^24^.

While plasma from COVID patients revealed an elevated mean shear and longitudinal viscosity compared to that of healthy persons (Supplementary Table 3, Supplementary Fig 2), patient-patient variability would limit its stand-alone diagnostic value on an individual basis. We thus in each case focus on the temperature-scaling behavior between 36-40.5°C (Fig 2A & 2B). We note that the propagated uncertainties in the presented values are smaller than the symbol size (see Methods, Supplementary Fig 3). Plotting the shear viscosity measured using a rolling-ball viscometer, as a function of the BLS longitudinal viscosity for healthy and COVID-19 patient plasma at different temperatures (Fig 2C), it is apparent that the two viscosities follow a similar trend (*inset* Fig 2C, Supplementary Fig 2B).

**Fig 2.**
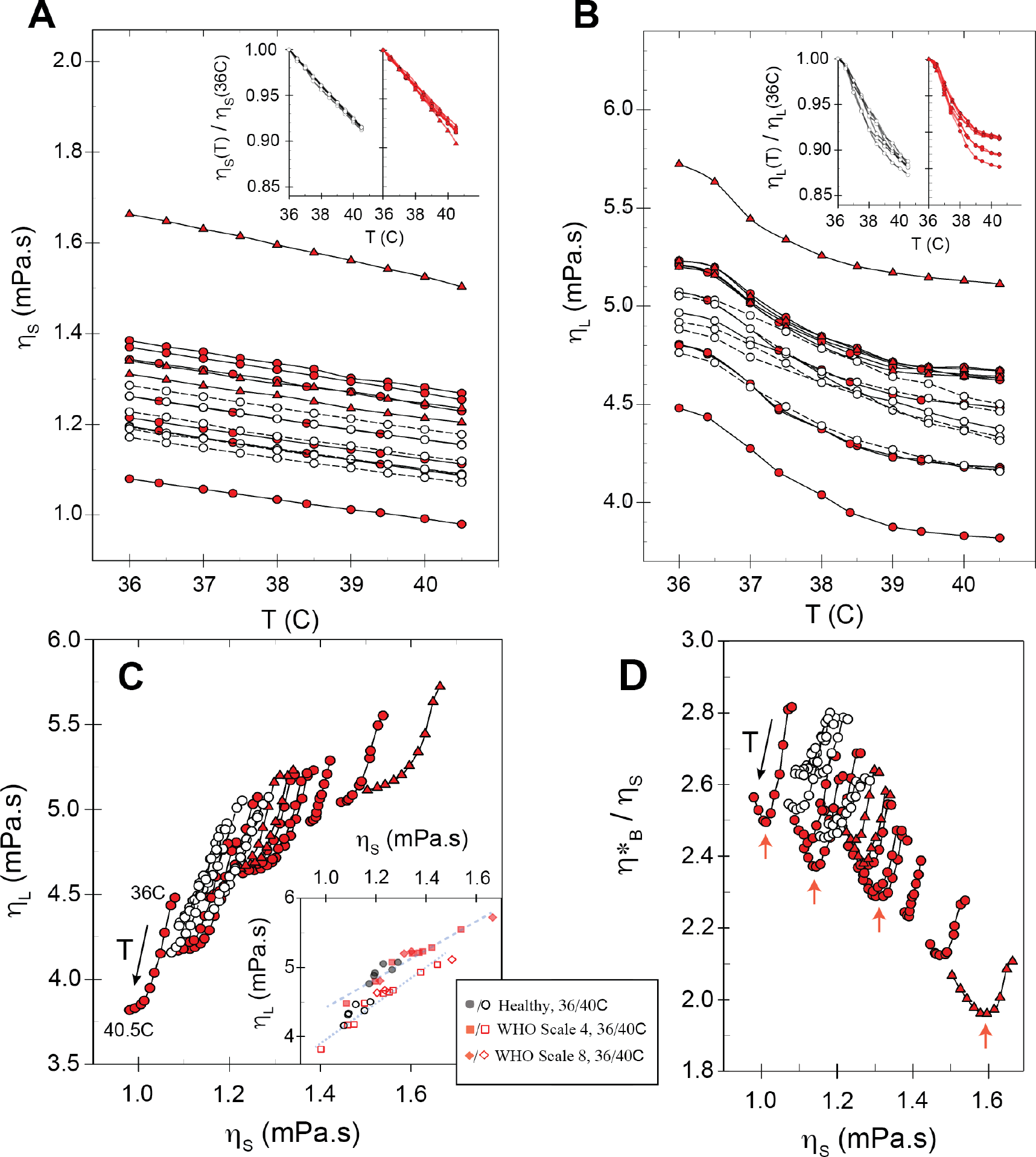
Temperature dependence of shear and longitudinal viscosities. **(A)** Temperature dependence of viscometer measured plasma shear viscosity *η*_&_. ∘ =healthy, 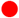 = COVID (WHO scale 4), 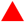 (WHO scale 8). Inset: *η*_′_ normalized to value at T=36C. **(B)** BLS measured longitudinal viscosity *η*_L_ for same samples as (A). *Inset: η*_L_ normalized to value at T=36C. **(C)** *η*_L_ plotted as a function of *η*_&_. *Inset:* same data shown for T=40.5C (solid symbols) and T=36C (open symbols), showing a common trend at both temperatures (See also Supplementary Fig. 2B). **(D)** Ratio of the effective bulk viscosity to shear viscosity as a function of shear viscosity (see Main Text). Red arrows indicate a minimum that occurs in most COVID patient plasma samples at a temperature of 38-39C.

**Fig 3.**
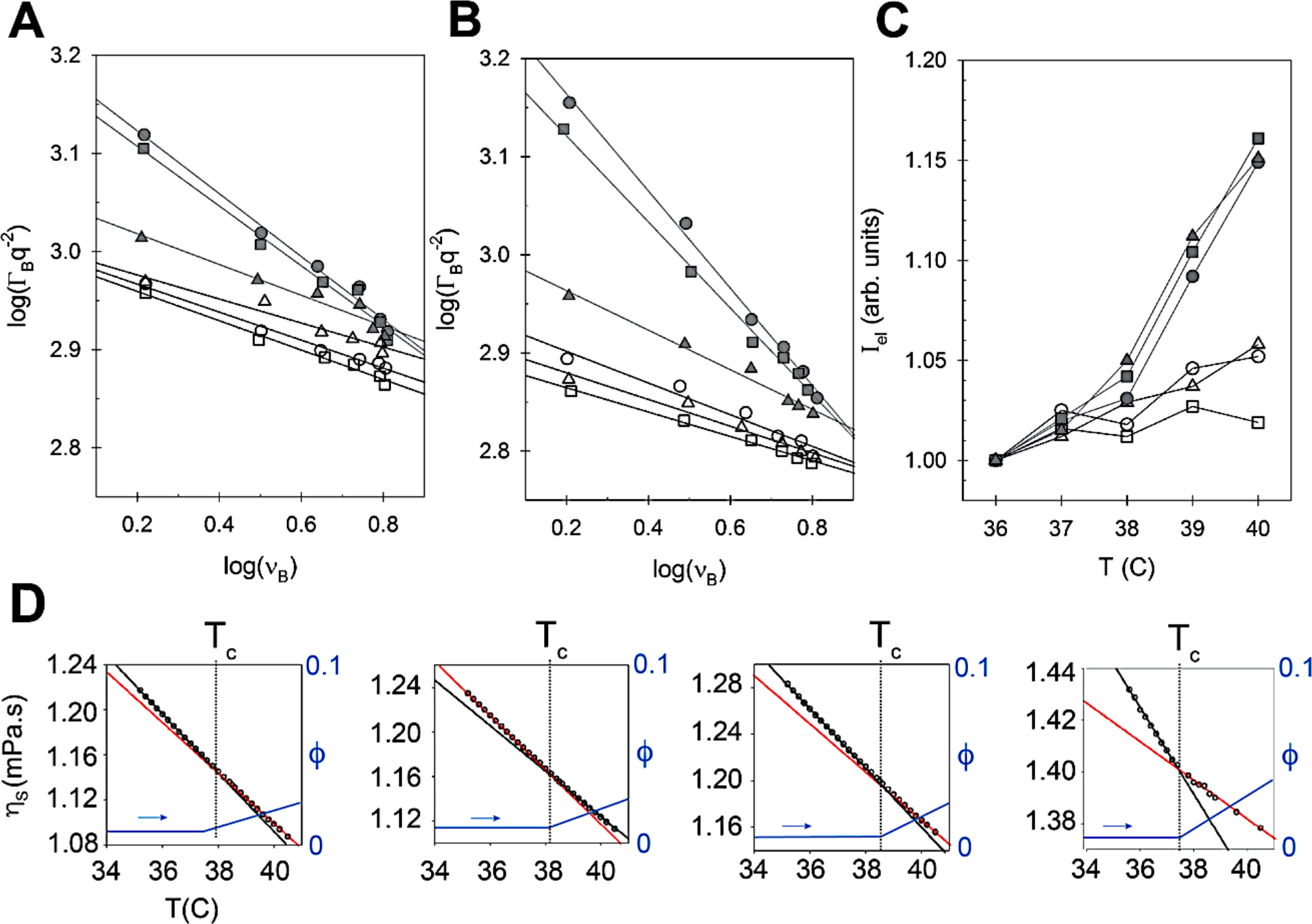
Anomalous dispersion, elastic scattering, and viscosity temperature scaling in COVID patient plasma. **(A)** Plot of log (Γ_*L*_*q*^(*s*^) as a function of log *v*_*L*_ for different scattering wavevectors (*q*) for healthy and COVID scale 8 patient plasma samples (open symbols = healthy; solid symbols = COVID WHO scale 8 patient samples) at T=36C. The slopes are given by δ = *b* − 2, where *b* is the frequency exponent of the attenuation coefficient (see Main Text). Fitted slopes of the healthy plasma samples are -0.143, -0.150 and -0.123, whereas those of COVID patient samples are -0.321, -0.305 and -0.157, consistent with a larger deviation from a pure fluid behavior. **(B)** Same measurements performed at T=39C (slope = -0.162, -0.136 and -0.124 for healthy patient samples and -0.496, -0.440 and -0.202 for COVID WHO scale 8 patient samples). **(C)** Intensity of elastic scattering peak (normalized to value at T=36C) as a function of temperature for the same samples. **(D)** Temperature dependent shear viscosity (*η*_&_) of four COVID WHO scale 8 patient samples showing a discontinuity in the slope at ≈38C. Also shown are suspensionfraction concentrations (*ϕ*) that could result in the observed *η*_′_ temperature dependence (see Main text).

To gain insight into the temperature scaling of *η*_*b*_ compared to *η*_*s*_, we can attempt to disentangle the shear and bulk contributions using Eqn. 1. In light of the non-Newtonian nature of plasma^2^, and *η*_*L &*_ *η*_*s*_ being measured at very different frequencies (see also Fig 1B), the calculated *η*_*b*_ will not yield the true bulk viscosity and will be referred to as the *effective bulk viscosity* 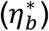 . Plotting 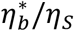 as a function of *η*_*s*_ we observe a distinct scaling behavior for the case of COVID-19 WHO scale 4 and 8 patient plasma (Fig 2D), which suggest that as the temperature increases (or *η*_*s*_ decreases) 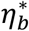 saturates faster in these samples.

We can probe the frequency-scaling over a narrow frequency-range, via angle-resolved BLS measurements, by tuning the measured scattering wavevector (and thereby the frequency). This is possible using a high Numerical Aperture (NA) objective and spatially masking different segments of the detection path^25^ (Methods). Doing so, we perform measurements on 3 healthy and 3 WHO scale 8 COVID-patient plasma samples. If at the probed frequency (*v* = *v*_*B*_) we are exclusively sensitive to fast solvent relaxation processes, the longitudinal acoustic attenuation coefficient would scale as *α*_*L*_ *∝ v*^*b*^ (and the longitudinal loss modulus as *M*^′^ *∝ v*^b−1^) with *b* = 2. Since *M*^′^ *∝ q*^−2^*v*_*B*_Γ_*B*_, the linewidth would scale with frequency as 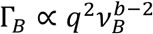, where 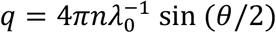, *n* is the refractive index, and *θ* the scattering angle (180° for backscattering). A plot of log (Γ_*B*_*q*^−2^) as a function of log *v*_*B*_ would thus have a slope of δ = *b* − 2 and reveal deviations from a quadratic frequency scaling of *α*_*L*_. This is shown for the 6 samples at 36°C in Fig 3A. Both healthy and WHO scale 8 COVID-patient plasma exhibit a negative slope (δ < 1), with two of the 3 WHO scale 8 COVID patient samples having a steeper slope, indicative of a larger deviation from quadratic frequency scaling. Performing the same measurements at 39C (Fig 3B) shows this slope to be notably more pronounced, corresponding to a decrease in *b*. This may suggest the additional presence or increased strength of slower relaxation process(es) in WHO scale 8 COVID samples that is more pronounced at elevated temperatures (*viz*. Fig 1B inset). Such a change could result from the increased presence of solid-like suspensions (proteins or protein aggregates), which have characteristic relaxation times on the ∼kHz scale^2^.

The presence of a suspension may also manifest itself by an increase in the elastic scattering. Elastic scattering is associated with thermally diffusive modes (Rayleigh scattering) and other collective relaxation processes, is however usually dominated by specular reflections in the optical setup and Tyndall scattering in the sample^26^. While specular reflection in the optical setup is sample independent, Tyndall scattering can be expected to increase with the presence of a solid-like suspension or aggregate. Plotting the normalized elastic scattering intensity as a function of temperature (Fig 3C) reveals that at elevated temperatures (above ≈38C) there is a notable increase in the elastic back-scattering for the WHO scale 8 COVID patient samples, consistent with an increase in solid-like constituents at elevated temperatures.

Interestingly, a closer inspection of the temperature dependence of the shear viscosity of COVID WHO scale 8 plasma samples exhibit a *kink* in the temperature-scaling at around 38-39C, above which the slope *dη*_*s*_/*dT* is smaller (Fig 3D), consistent with a change in the physical properties occurring at these temperatures. This appears to also be more significant for samples with a higher absolute shear viscosity (Fig 3D).

To investigate whether any sizable suspensions might be present in the COVID-19 patient plasma we perform fast spatial BLS scans of healthy and COVID-19 patient samples (Methods). We observe a spatial heterogeneity in several of the latter, marked by a punctuated presence of distinct viscoelastic features, that become more pronounced at elevated temperatures. This is shown in Fig 4A and 4B, where we plot the loss tangent defined as tan 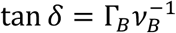 which to a good approximation is independent of the refractive index and mass density and thereby represents a change in the viscoelastic properties that does not assume values of these. We note that due to diffusion and finite voxel dwell times in our scans, these spatial maps are not representative of the size of such heterogeneities and by extension the values will not be a true quantitative measure of their viscoelastic properties (see Methods).

**Fig 4.**
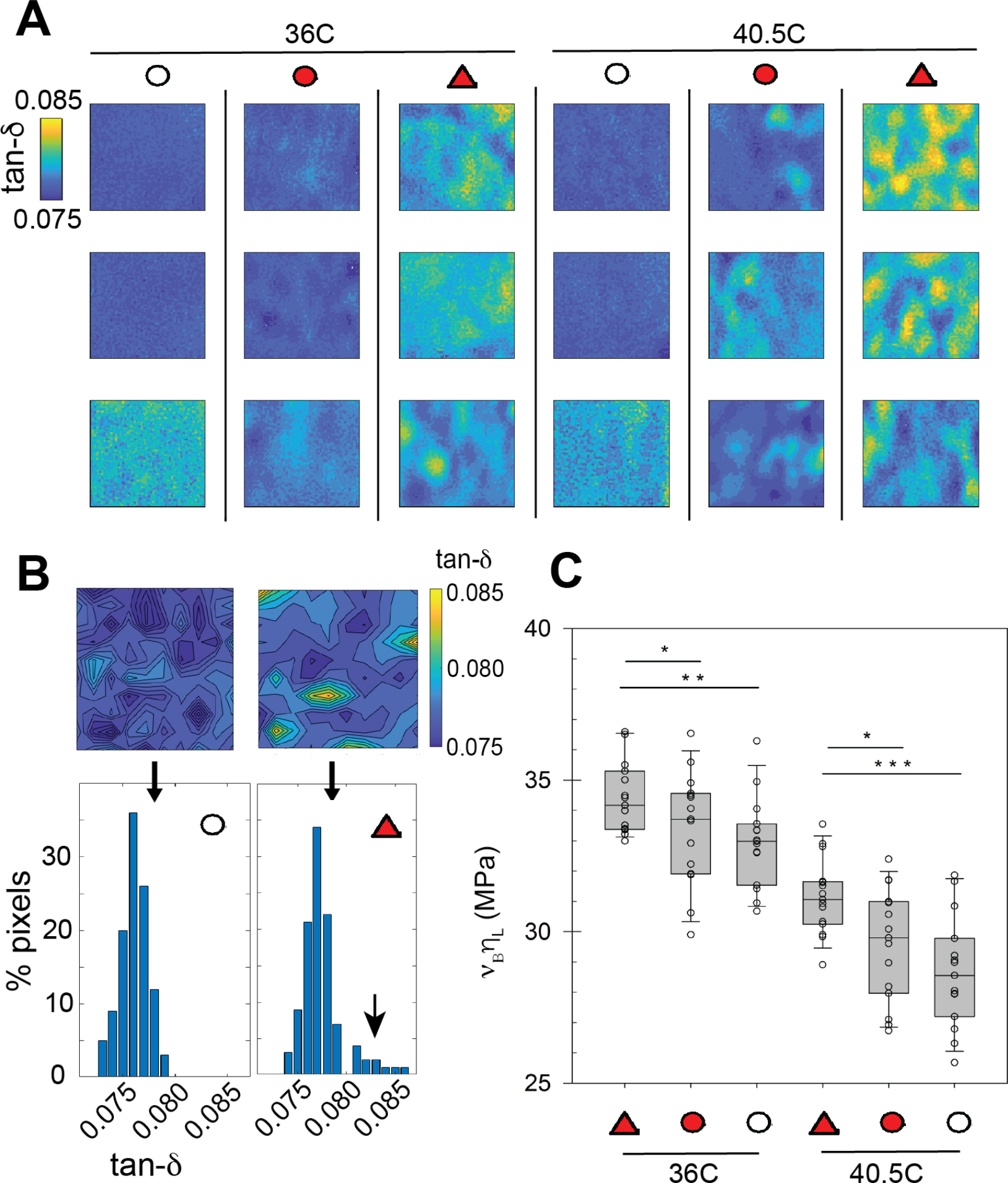
**(A)** Confocal BLS spatial maps of the loss tangent 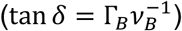 in 3 samples from healthy (∘), and COVID patients with WHO Scale 4 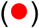 and 8 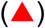, showing an increased heterogeneous viscoelastic landscape in COVID patients that is more pronounced at elevated temperatures. Scan in each case is 200x200μm. **(B)** Quantification of scans showing the presence of viscoelastically distinct regions in COVID patient samples (T=36). **(C)** Collated values of the product *v*_*L*_*η*_L_, which would increase with increasing suspension concentration (see Main Text), calculated as the average value from 1mm square spatial scans of 10 samples for each case. Boxes indicate interquartile ranges with median line, and whiskers are 5%/95% range. Statistics performed using unpaired *t*-test with Welch’s correction (*p<0.05, **p<0.01, ***p<0.001).

The data presented in Fig 4B are averages of many individual measurements probing ∼μm-mm sized volumes (Methods). As such, the effective measured complex longitudinal moduli of plasma containing solid -like constituents with a distinct effective modulus *M*_a_, may be written as a weighted average: *M* = (1 − ϕ)*M*_/_ + ϕ*M*_a_, where ϕ is the solid-constituent volume fraction and *M*_*p*_ the effective modulus of the plasma (water, electrolytes, non-aggregated proteins, etc.) in their absence. This is simply a statistical average (independent of the mixing-law^6,7,27^), and values of *M*_a_ and *M*_/_ should be seen as effective rather than true values for the constituents. It follows however that at a given temperature the spatial average < *η*_*L*_*v*_*B*_ > will scale linearly with respect to *ϕ* as: 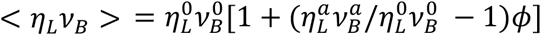, where superscripts “*a*” and “0” denote the respective values for the solid-constituents and for the plasma in their absence. Collated values of < *η*_*L*_*v*_*B*_ > for COVID-19 (WHO scale 8) and healthy patient plasma are shown in Fig 4C, revealing that <*η*_*L*_*v*_*B*_ > is notably larger for plasma from COVID WHO scale 8 patients than both that of WHO scale 4 patients and healthy patients. Interestingly the difference seems to be more pronounced at higher temperatures, consistent with an increased solid-like constituent volume fraction at higher temperatures, as is suggested also by our observations for the frequency scaling (Fig 4A and 4B) and elastic scattering (Fig 4C) data.

To see if an increased solid fraction at higher temperatures would be consistent with the observed shear and bulk viscosity temperature scaling, we theoretically consider the effect of solid-like suspensions on the scaling of the two viscosities. The shear viscosity can to a first approximation be assumed to depend on a suspension concentration *ϕ* as^28^:

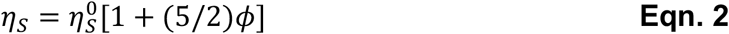

where 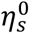 is the shear viscosity in the absence of the suspension. The precise functional dependence will in general be more complicated, and depend on the suspension size, properties of the solvent, maximum volume fraction, *etc*.^28^, however for small *ϕ* this (Einstein approximation) is a reasonable functional approximation. Assuming the contribution from suspensions only becomes significant in COVID-patient samples, we take sample averaged values of the shear viscosity from healthy plasma samples for 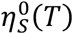 and calculate *η*_*s*_(*T, ϕ*) for the case of different *ϕ*. Based on the monotonic increase in the elastic scattering cross-section for COVID patient samples above 38-39C (Fig 3C) we as a first approximation take:

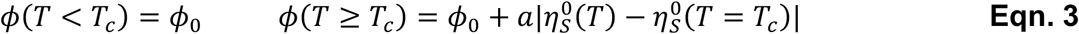

Here the parameter *a* will determine the rate of *suspension change* with respect to temperature. The scaling with respect to 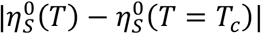 is motivated by the observation that the change in *dη*_*s*_/*dT* is more pronounced in samples with a stronger shear viscosity temperature dependence (Fig 3D). We find that this temperature dependence can qualitatively describe the kink in the shear viscosity observed at 38-39C (Fig 3D).

The effect of a suspension on the bulk viscosity, while the subject of several theoretical investigations, is less trivial and will depend on their bulk properties relative to the solvent^29,30^. For solid-like inclusions we may as a first approximation write^29^:

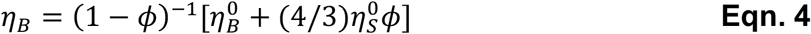

where 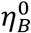 is the bulk viscosity in the absence of solid-like inclusions. Again, taking sample averaged values of the effective bulk viscosity from healthy plasma samples for 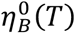 and using the same temperature dependent suspension volume fraction as for the shear viscosity (Eqn. 3) we can calculate *η*_*B*_(*T*) for given values of *a*. The temperature scaling of the shear and bulk viscosity calculated from Eqn. 1-4 for different values of *a* are shown in Fig 5A and Fig 5B, where it can be seen to predict the discontinuity on the temperature scaling in *η*_*s*_(*T*) (inset of Fig 1A, and Fig 3D) as well as the saturation in *η*_*B*_(*T*) (inset of Fig 1B, and Fig 1D) observed in COVID-19 patient plasma samples.

**Fig 5.**
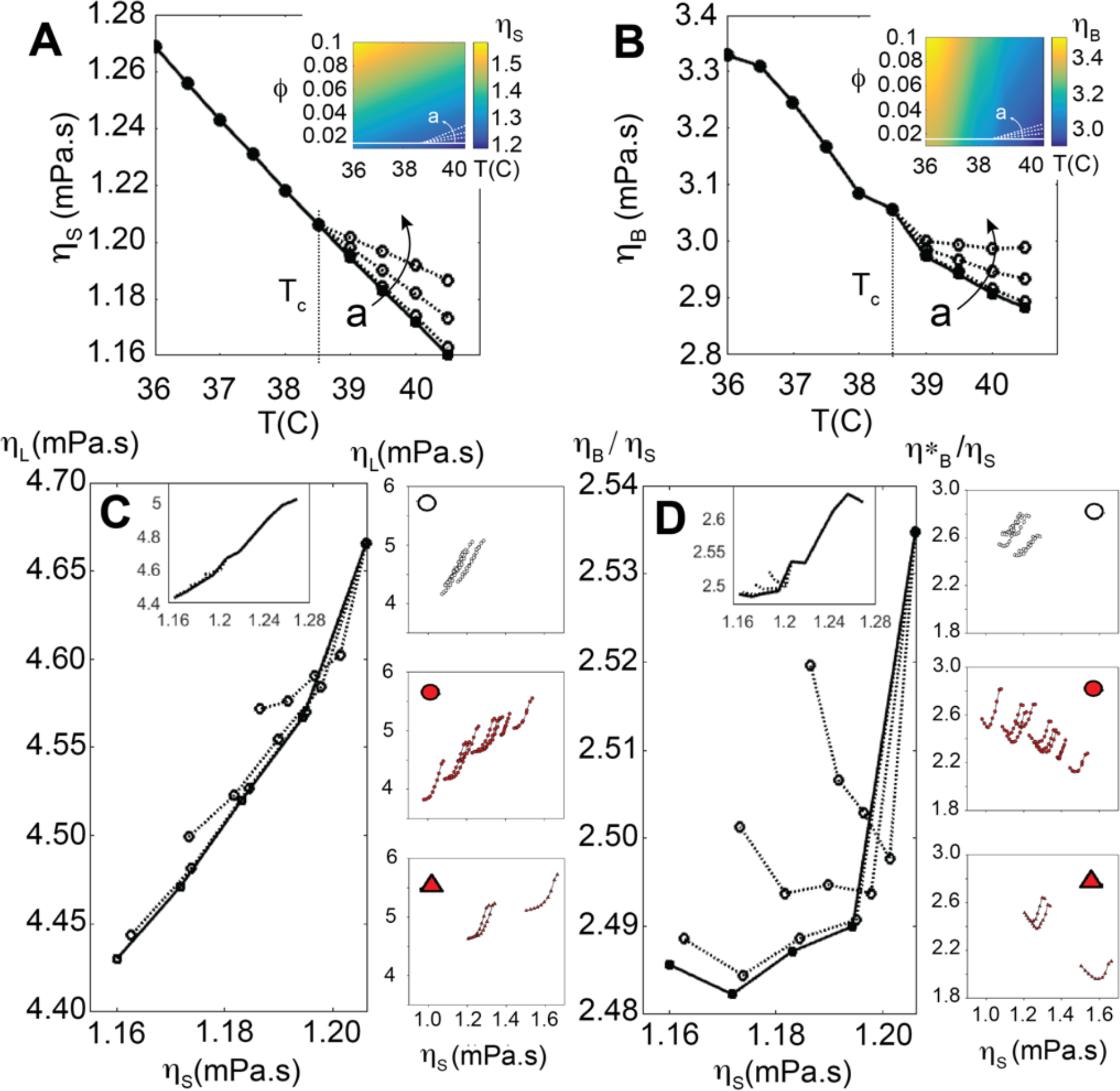
Scaling of shear, bulk and longitudinal viscosity for increasing suspension concentrations. **(A) & (B)**: Shear and Bulk viscosity calculated using Eqn 2 and 4 with the suspension concentration *ϕ* give by Eqn 3, shown for increasing values of the parameter *a* (see Main text). *Inset:* Heat map of respective plots showing trajectory of *ϕ*. **(C) & (D)**: Calculated *η*_L_ and 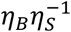 plotted as a function of *η*_′_ for increasing values of *a*. Also shown are corresponding experimental results for healthy (∘), and COVID patients with WHO Scale 4 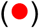 and 8 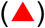.

## Discussion

We present a study of the BLS-measured longitudinal viscosity *η*_*L*_ of COVID-19 patient blood plasma with different disease outcomes, and compared these to the viscometer-measured shear viscosity *η*_#_. We find *η*_*L*_ can serve as a proxy for the viscometer-measured *η*_#_ but observe no clear sample independent proportionality between the two, consistent with *η*_*B*_ representing an independent material property. Our results suggest that BLS may be used to assess increased PV however quantitative comparisons to conventional PV measurements should be approached with caution. Our results also show that although increased *η*_*L*_ and *η*_#_ may be indicative of COVID infection severity, patient-patient variability may limit their stand-alone diagnostic usefulness.

Interestingly we find that at temperatures above ∼38°C *η*_*L*_ exhibits a saturation that is more pronounced in severe COVID-patient samples, which is also accompanied by a kink in the temperature scaling of *η*_#_. A simple theoretical description of the scaling of *η*_*L*_ and *η*_*s*_ with respect to suspension concentration, suggests our observations may be described by an increase in suspension concentration above ∼38°C in severe COVID-patient samples. This is supported by BLS confocal maps which reveal >micron-sized spatial heterogeneities in COVID-patient plasma absent in healthy patient samples, that increase at elevated temperatures, and are accompanied by an increase in the elastic scattering cross-section. Measurements of the frequency-scaling of *η*_*L*_ by angle-resolved BLS also suggest more pronounced >picosecond relaxation processes in COVID patient samples at elevated temperatures, consistent with an increased suspension concentration.

Though a significant increase of FDPs can conceivably result in viscoelastic changes, these in of themselves would be too small in size to explain the observed spatial heterogeneities, and would not explain the lack of any significant hysteresis in up-down temperature sweeps observed (which also rule out the denaturing of proteins). Our observations are somewhat surprising given that our samples were collected and stored in anticoagulation tubes (Methods), although reports have hinted at the reduced effectiveness of anticoagulants in COVID-19 patients^31,32^. While ultrastructural and biochemical studies on suspensions is still needed, one can speculate whether these might be related to crosslinked microfibrin/fibrin microclots reported to exist in COVID patient samples^33^. Should this be the case this would suggest that the balance between increased fibrin crosslinking (clot formation) and the increased dissociation rate with increasing temperature^34^ is perturbed in COVID patient plasma. To this end BLS temperature-scaling measurements, in addition to potentially providing a means of assessing COVID severity, could also serve as a useful tool for exploring the viability of different drugs in mitigating suspension formation. Given the persistence of crosslinked fibrin aggregates and microclots also in Long-COVID patient plasma^33^, a means of quantitatively assessing their abundance can be of prognostic and diagnostic value. *Longitudinal rheology*, may be well suited for this, owing to its acute sensitivity to suspension properties.^7^

While the last decade has seen significant technological strides in BLS spectrometers for life-science applications^35^, the field has largely focused on the more easily obtainable longitudinal storage modulus. There is however growing awareness that changes in the longitudinal viscosity might offer a clearer indicator of structural-physical changes in soft matter^27,36^. Unlike the longitudinal storage modulus, which will in pure liquids be given by the bulk modulus (and often dominated by this in soft matter^37^), the contribution of shear and bulk components to *η*_*L*_ are on roughly equal footing: with values of *η*_*b*_ for different fluids of the same order of magnitude as *η*_#_9,38. BLS has several desirable features including its non-destructive all-optical label-free nature, fast acquisition times (<second), high sensitivity and repeatability, and small required sample volumes (∼microliter). There are several limitations--Supplementary Text. To serve as a rapid diagnostic/prognostic tool, one is primarily limited by the means of separating plasma from whole blood. For this the combination with existing techniques used for separating erythrocytes from plasma (plasmapheresis), or custom microfluidic devices and ones employing standing acoustic waves^39^, may be envisioned for real-time monitoring of the longitudinal plasma viscosity (Supplementary Text). Together with progress in BLS spectrometer miniaturization^40^ and compact BLS-probes^41^, this may allow for the realization of compact instruments for routine point-of-care/ICU applications to asses disease criticality.

## Data Availability

Data for this study are available in the Main Text or Supplementary Materials, and where not the case, will be made available upon publication via a DOI repository such as Zenodo.

## Acknowledgments

This work was supported by funding from the Austrian Science Fund-FWF (Grant No. P34783). No funding sources had any role in the writing of the manuscript or the decision to submit.

## Author contributions

Conceptualization & project administration: AZ, MF, KE. Methodology: JI, TS, JHA, KE. Investigation & visualization: JI, TS, KE. Funding acquisition: WJW, KE. Supervision: AZ, MF, KE. Writing – original draft: KE. Writing – review & editing: All Authors

## Competing interests

All authors declare that they have no competing interests.

## Code availability

Code used for analysis of data that has not already been published elsewhere will be made available upon publication via a DOI repository such as Zenodo.

## Supplementary Materials

Supplementary Table 1-3, Supplementary Figures 1-3, Supplementary Text.

## Methods

### Blood plasma

Patient age ranged from 32-78 years (mean 61 years, median 62 years), with a male to female ratio of approximately 2:1 (Supplementary Table 2). Of the 39 COVID-patients, 18 were WHO scale 4 (no oxygen therapy), 2 were WHO scale 5 (oxygen by mask), and 19 were WHO scale 8 (intubated/mechanical ventilation)^23^. In addition to age and gender, Body Mass Index (BMI) on admittance to the hospital, the 28 and 90 day mortality, blood-pressure/hypertension, and underlying conditions including diabetes, coronary heart disease, chronic heart failure, chronic renal failure, immunosuppression, thyroid disorders, chronic lung diseases, active hematological diseases, active cancers, as well as whether they were active smokers, were recorded. A significant larger fraction of WHO scale 8 patients than WHO scale 4-5 patients had one or more of these underlying conditions (84% *vs*. 35%), and a notably larger fraction also exhibited hypertension (74% *vs*.15%) (Supplementary Table 2). Blood samples from patients were collected in EDTA tubes and immediately centrifuged at 3000 rpm for 15min. The plasma was subsequently transferred to Eppendorf transport tubes for BLS and viscometer measurements (see below).

### BLS microspectroscopy

The BLS spectrometer used was a dual (cross-dispersion) Virtual Imaged Phased Array (VIPA) based imaging spectrometer^42^ similar in design to that previously described^43^. Notable differences where that it was designed to operate at 660nm (laser source: “Flamenco” SLM 660nm 200mW by Cobolt/Huber Photonics, Germany). The choice of using 660nm vs. the more common 532nm for this setup was based on the lower absorption of blood plasma at these wavelengths^44^ that would result in reduced unwanted laser heating and potential protein photodamage. The two VIPAs (Light Machinery, Canada) each had a free spectral range of 15GHz at 660nm, with reflective and anti-reflection coatings on the entrance window optimized for this wavelength. The spectra was imaged using an sCMOS (Edge 4.0, PCO, Germany) prior to which a Lyott stop was included^45^ to serve as a low pass filter for removing interference scattering effects from the sample and optical setup. Owing to the reasonable transparency of the plasma no extra elastic suppression was required, beyond the spatial masking of the elastic scattering by two slits at intermediate image planes. In the final spectral projection the magnification was such that the Brillouin peaks occupied around 5-10 camera pixels (example spectra of BLS peaks are shown in Supplementary Figure 3A).

The microscope setup itself consisted of a custom-built inverted microscope with a 2.5cm x 2.5cm motor translations stage (ASI, Canada) and temperature controlled sample box (Ibidi, Germany). Measurements were performed in backscattering geometry with a low NA objective (0.13 NA 4X, RMS4X-PF, Olympus, Japan) to minimize line broadening^46^. The polarization of the laser light was adjusted with a half waveplate, expanded to ∼18mm with a relay lens system (to overfill the back aperture of the objective lens), and coupled into the optical setup via a Polarizing Beam Splitter (PBS), with a quarter waveplate prior to the objective lens. As such the sample was probed with circularly polarized light, but given that in the backscattering geometry and in liquids we would only measure longitudinal modes this is not an issue. The scattered light was focused using a similar 0.13 NA objective into a single mode fiber (which served as a pinhole of effective size 1-2 Airy Units) that ran to the BLS spectrometer. By adjusting the half-waveplate in front of the laser the light could alternatively be preferentially coupled to a calibration sample in a cuvette (water and PMMA) behind the PBS, that was used for spectral registration prior to each imaging session as previously described^41^. All components and acquisition were controlled by a LabView based script developed by THATec Innovations (Germany); while spectral registration, deconvolution and data analysis was performed in Matlab (Mathworks, Germany) as previously described^43^. Deconvolution to extract the true linewidth was performed by noting that in Fourier space the measured signal is simply the product of the spectral response function and the true signal. For the former we used a measurement of the elastic scattered signal made before each imaging session obtained by placing a mirror flat in place of the sample, attenuating the laser with neutral density filters, and opening the masking slits in the spectrometer. We note that deconvolution appeared to reduce the linewidth by a small almost constant amount and had no effect on any of the trends or qualitative conclusions presented.

For each measurement ∼750μL of plasma from the EDTA storage tubes was pipetted onto a glass bottom dish (Matek, Slovania), after shaking the former for ∼20 seconds with a vortex mixer. We note that given the probed confocal volume this volume of plasma is much more than is needed (and one could theoretically work with microliter sized volumes). The glass bottom dish was subsequently placed in the temperature enclosure (set at 36C) on the microscope stage (Ibidi, Germany). We waited for 15min for the sample temperature to equilibrate (during which spectral registration measurements were performed), before performing the first measurements. The temperature of the sample was monitored by means of a needle temperature probe in contact with the sample with an accuracy of <0.1C prior to each measurement, and the temperature and humidity of the sample enclosure via an additional temperature probe. The upper temperature limit of 40.5C for the temperature sweeps was set based on the observed non-reversibility of properties above these temperatures (presumably due to protein degradations).

For measurements we focused the laser approximately 100μm above the coverslip surface (such that the probed confocal volume is fully within the plasma). Measurements consisted of a 4x4 lateral square grid scan with step-spacing 250μm in approximately the center of the sample and was controlled by the stepper motor. The summed spectra of these 16 points were used for further analysis. In each case the laser power at the sample was approximately 40mW and the exposure time 250ms.

The Brillouin frequency shift (*v*_*B*_) and line width (Γ_*B*_) were obtained by fitting the measured spectrum *I*(*v*), after spectral registration and deconvolution, with a Damped Harmonic Oscillator (DHO):

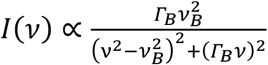

The longitudinal viscosity (*η*_*L*_) was calculated from the Brillouin linewidth Γ_*B*_ via:

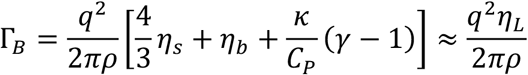

Here *ρ* is the density, *κ* is the thermal compressibility, and *γ* = *C*_*p*_⁄*C*_v_, where *C*_*p*_ and *C*_v_ are the specific heat at constant pressure and volume respectively. Since to a good approximation we can assume *C*_*p*_ = *C*_v_ in plasma, the second term in the parenthesis disappears and the line width becomes directly proportional to the longitudinal viscosity. Here 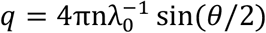, where λ_0_ = 660nm is the free-space probing wavelength, *n* is the refractive index, and *θ* here is the scattering angle. In the back-scattering geometry with low numerical aperture probe/detection we can assume sin(*θ*/2) → 1. The refractive index of aliquots of the same samples were measured using an Abbe refractometer and ranged from 1.338 to 1.348. Measurements of the same samples at 40.5C resulted in a change of <1% for a given samples and as such the temperature dependence could be ignored for our calculations.

The loss tangent was calculated via:

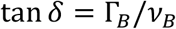

The precision with which the BLS-shift and linewidth is measured is estimated by performing a 10x10 grid scan of 100 points (5μm step size) of distilled water, and calculating the standard deviation of the obtained values for the frequency shift and linewidth. This is shown for different exposure times in Supplementary Figure 3, where we see for the employed laser power (40mW) and exposure time (250ms) the precision in *v*_*B*_ and Γ_*B*_ are 3.1MHz and 6.1MHz. Propagating this uncertainty for the derived parameters in Fig. 2-4 would result in error bars smaller than the size of the respective symbols. Decreasing the laser power by 25% and 50% decreased the SNR correspondingly (and thereby reduced the precision), as is shown in Supplementary Figure 3A. Here the SNR was calculated as the average maximum intensity of the BLS peak of the 100 point scan, divided by the standard deviation of these maxima. Importantly, decreasing the laser intensity negligibly affected the mean peak position and width (Supplementary Figure 3B), which remained in agreement within experimental uncertainties, and suggesting that the employed laser intensity does not affect our results. This also suggests there are no significant perturbations due to local heating/damage of the sample which would potentially increase the BLS shift and decrease the linewidth.

Confocal BLS spatial maps (Fig 4) were performed using a different dual-VIPA Brillouin microscope of a conceptually similar optical design but optimized for 532nm with an FSR of 30GHz, equipped with a Piezo and Motor scanning stage and sample heater, as previously described^41,43^. This was additionally equipped with an Apodization filter^35^, Lyot Stop^45^, and Iodine absorption cell^47^. The pixel dwell time was 25ms, and the excitation power at the sample was 30-35mW. Measurements were performed by continuously sweeping the scanning stage (linear ramp) during acquisition and as such the value for each pixel corresponds to the average of the distance between adjacent pixels which was 2μm. For this reason the maps in Fig 4 can also not be quantitatively interpreted to represent the heterogeneity on spatial scales below this. The variable power was the result of the laser current being modulated to lock the laser frequency to an absorption line of the gas cell via a feedback loop^41^ which differed slightly between consecutive measurements. The precision of these measurements for both the fitted peak position and linewidth was < 5 MHz (based on a 100 pixel scan of distilled water as described above).

#### Angle-resolved BLS measurements

Angle-resolved BLS measurements (Fig 3A and 3B) were performed using the same setup as for the BLS setup operating at 532nm with the following modifications. The objective was replaced with a high numerical (NA) objective (1.4 NA immersion 60X Olympus). The relay lens system used to expand the laser beam to fill the back aperture of the objective was modified such that the collimated beam incident on the back aperture had a diameter of about 1mm. This corresponded to an effective probing NA just under 0.1. On the detection side we placed an annular mask (406 aluminum coated quartz, custom made by Photo-Sciences Inc., USA) which consisted of angular rings with different diameters^48^. The mask itself contained >20 different angular ring diameters and by selectively placing different angular ring diameter masks in the light path we can probe the BLS scattering at different scattering wavevectors (with a detection NA of approximately 0.1). In our experiments we used 4 different ring diameters. The scattering angles in the sample (and probed wavevector) could readily be calculated from the ring diameter and refractive index of the plasma sample. The acquisition time for these measurements was 5-10 seconds, which was required to obtain a satisfactory SNR of the BLS peaks. Analyzed spectra was taken as the sum of a 10 x 10 grid scan, and spectrally deconvolved with the instrument spectral response in the same manner as with the conventional BLS measurements.

#### Elastic scattering measurements

The elastic scattering measurements (Fig 3C) were performed in the same manner as the BLS measurements but with the intermediate imaging plane masking slits in the spectrometer open, and the signal attenuated by a neutral density filter placed immediately prior to coupling into the spectrometer (such that the elastic peaks were only ∼50% saturated on the spectrometer camera). The intensity (in arbitrary units) was taken as the average of the peak maximum of two elastic scattering peaks from adjacent orders. The elastic scattering from specular reflections in the spectrometer and Tyndall scattering were assumed to dominate, given that the contribution from other collective modes are much weaker.^49^ Also the Rayleigh scattering can be assumed to be comparatively small as it will to a first approximation scale as 1 − *C*:/*C*_9_, where *C*: and *C*_9_ are the specific heats at constant volume and pressure, which are roughly comparable. Since the contribution from specular reflection inside the spectrometer is sample independent, changes in the elastic scattering intensity can be attributed to changes in the Tyndall scattering. Extra care was taken to assure that there was no additional contamination on the sample mounting side, with new sterile individually packaged glass bottom dishes used for each measurement. As such changes in the scattering can be attributed to changes in sample optical inhomogeneities.

### Shear viscosity measurements

Temperature dependent measurements of the shear viscosity were performed on aliquots of the same plasma samples in parallel to or immediately following BLS measurements. For these a commercial rolling-ball shear viscometer (Lovis 2000, Anton Paar, Austria) calibrated to ASTM D2162/ UKAS ISO 17025 /ISO 17034 certified water viscosity standard, was used. Routine annual calibration of the instrument by the manufacturer assured that the measurements can be considered reliable. Following established manufacturer-provided protocols, automated programmed temperature-ramps 36C-40.5C-36C were performed, with for each temperature step the averages of two iterations taken. The respective uncertainties were smaller than the symbol sizes in Fig 2 and 3.

Modeling of the suspension dependence of the shear and longitudinal viscosity was performed using Matlab (Mathwork, Germany), and all statistical analysis was performed using Sigmaplot 13.0 (Sysworks, Germany).

The study was performed in accordance with the recommendations of the Declaration of Helsinki. Experimental protocols were approved by the Ethics Committee of the Austrian Federal Agency for Safety in Health Care - BASG (ECS 1372/2021, Ethics votum:EK-21-068-VK, BASG Operation number:14043408) and written informed consent was obtained from the patients.

## Notes

### Competing Interest Statement

The authors have declared no competing interest.

### Author Declarations

Ethics Committee of the Austrian Federal Agency for Safety in Health Care (BASG) gave ethical approval for this work.

