## Supplementary Material for "Longitudinal viscosity of blood plasma for rapid COVID-19 prognostics"

This file includes:

**Supplementary Tables 1-3**

**Supplementary Figures 1-3**

**Supplementary Text**

#### Supplementary Tables

| Technique | Measurable parameters* | Frequency | Acoustic length-scales [wavelength / decay-length] | minimum sample volume** | Typical measurement duration | Precision*** [acoustic speed / attenuation] | Readout mode |
|---|---|---|---|---|---|---|---|
| Brillouin light scattering spectroscopy | $M', M''$, $\eta_L, \alpha_L$ | ~1-10 GHz | ~100 nm / ~1μm | < 100 μL | ms - sec | <0.1% / < 1% | *Optical* (inelastic scattering spectrum) |
| Acoustic spectroscopy | $M', M''$, $\eta_L, \alpha_L$ | ~1-10 MHz | ~100μm / ~1mm | > 15 mL | ~sec | 0.03% / 0.7% | *Mechanical-electrical* (transducer perturbation) |

**Supplementary Table 1: Comparison of Brillouin Light Scattering spectroscopy and Acoustic spectroscopy.** * = Both Brillouin light scattering spectroscopy and Acoustic spectroscopy can also extract the shear modulus and shear viscosity in non-Newtonian fluids, however these are significantly more challenging to obtain. ** = To assure temperature stability the sample volume may need to be larger, however, this would in itself be independent of the technique used. *** For measuring water at ~35C. As described in Methods for Brillouin Light Scattering spectroscopy (see also Supplementary Fig 3), and in Ref[9] for Acoustic spectroscopy.

| WHO scale | O2 | NIV | IV | 28 day mortality | 90 day mortality | hyper-tension | obesity (*BMI*) | diabetes | coronary artery disease | chronic heart failure | chronic renal failure | immuno-suppression | thyroid disorder | chronic lung disease | current active smoking | active cancer | hematological disease | |
|---|---|---|---|---|---|---|---|---|---|---|---|---|---|---|---|---|---|---|
| 4 | | | | | | ■ | ■ *28.0* | ■ | | | | | | | | | | |
| 5 | ■ | | | | | | *24.8* | ■ | | | | | | | | | | |
| 4 | | | | | | ■ | *22.0* | | | | | | | | | | | * |
| 4 | | | | | | | ■ *29.8* | | | | | | | | | | | * |
| 4 | | | | | | | ■ *30.8* | ■ | | | ■ | | | ■ | | | | |
| 4 | | | | | | | ■ *28.1* | | | | | | | | | | | |
| 4 | | | | ■ | ■ | | ■ *34.3* | ■ | | | | | | ■ | | | | |
| 4 | | | | | | | *25.0* | | | | | | | ■ | | | | |
| 4 | | | | | | | ■ *31.6* | | | | | | | ■ | | | | |
| 4 | | | | | | | ■ *33.1* | | | | | | | | | | | * |
| 4 | | | | | | | *24.9* | | | | | | | | | | | * |
| 4 | | | | | | | ■ *27.8* | | | | | | | | | | | * |
| 4 | | | | | | | *24.2* | | | | | | | | | | | * |
| 4 | | | | ■ | ■ | | ■ *28.0* | | | | | | | | | | | |
| 4 | | | | | | | ■ *27.3* | | | | | | | ■ | | | | |
| 5 | ■ | | | ■ | ■ | | *23.4* | | | | | | | | | | | |
| 4 | | | | | | | *24.0* | | | | | | | | | | | * |
| 4 | | | | | | | *25.0* | | | | | | | | | | | * |
| 4 | | | | | | | ■ *26.1* | | | | | | | | | | | * |
| 4 | | | | | | | *24.3* | | | | | | | | | | | |
| 8 | ■ | ■ | ■ | ■ | ■ | ■ | *26.0* | | | | ■ | | | | | | | |
| 8 | ■ | ■ | ■ | | ■ | | *25.0* | | | | | | | | | | | * |
| 8 | ■ | ■ | ■ | ■ | ■ | ■ | *29.1* | | ■ | ■ | | | | | ■ | | | |
| 8 | ■ | ■ | ■ | ■ | ■ | | *27.4* | | | | | | | | | | ■ | |
| 8 | ■ | ■ | ■ | ■ | ■ | ■ | ■ *31.3* | | | | | | | ■ | | | | |
| 8 | ■ | ■ | ■ | ■ | ■ | ■ | ■ *38.6* | ■ | | | ■ | | | | | | | |
| 8 | ■ | ■ | ■ | ■ | ■ | ■ | *26.2* | | | | | | | | | | | * |
| 8 | ■ | ■ | ■ | ■ | ■ | ■ | *26.2* | | | | | | | | | | | * |
| 8 | ■ | ■ | ■ | ■ | ■ | ■ | *29.8* | | | | | | | ■ | | | | |
| 8 | ■ | ■ | ■ | ■ | ■ | ■ | *29.1* | ■ | ■ | | | | | | | | | |
| 8 | ■ | ■ | ■ | ■ | ■ | ■ | ■ *43.0* | ■ | ■ | | | | | | | | | |
| 8 | ■ | ■ | ■ | ■ | ■ | ■ | *23.0* | ■ | ■ | ■ | ■ | | | | | | | |
| 8 | ■ | ■ | ■ | ■ | ■ | | ■ *36.4* | | | | | | | | ■ | | | |
| 8 | ■ | ■ | ■ | | | | ■ *40.0* | | | | | ■ | | | | | | |
| 8 | ■ | ■ | ■ | ■ | ■ | ■ | *25.7* | | | | | | | | | | ■ | |
| 8 | ■ | ■ | ■ | ■ | ■ | ■ | *21.0* | | | | ■ | ■ | | | | | | |
| 8 | ■ | ■ | ■ | ■ | ■ | ■ | ■ *49.6* | | | | | | ■ | ■ | ■ | | | |
| 8 | ■ | ■ | ■ | ■ | ■ | | *24.1* | | ■ | | | | | ■ | | | | |
| 8 | ■ | ■ | ■ | | | ■ | *28.0* | | ■ | | | | | | | | ■ | |

**Supplementary Table 2: WHO scale 4,5 and 8 COVID patient plasma considered for the study.** O2= supplied with oxygen, NIV = Non-Invasive Ventilation, and VI= Invasive ventilation. (In each case shaded black = affirmative). Shown also is patient mortality (black shaded = did not survive), and identified underlying health conditions (shaded = affirmative). “ * ” in the last column indicates samples where patients had none of the listed existing underlying conditions and which were further analyzed in our studies.

| | $\eta_S(36C)$ mPa.s | $\eta_S(40.5C)$ mPa.s | $\eta_L(36C)$ mPa.s | $\eta_L(40.5C)$ mPa.s |
|---|---|---|---|---|
| **Healthy** | 1.22 (± 0.02) | 1.12 (± 0.02) | 4.94 (± 0.05) | 4.36 (± 0.05) |
| **COVID *(WHO 4)*** | 1.31 (± 0.05) | 1.21 (± 0.05) | 5.07 (± 0.11) | 4.51 (± 0.13) |
| **COVID *(WHO 8)*** | 1.44 (± 0.11) | 1.31 (± 0.09) | 5.38 (± 0.17) | 4.81 (± 0.15) |

**Supplementary Table 3: Shear and Longitudinal viscosity of healthy and COVID-19 patient plasma.** Mean values of shear and longitudinal viscosity ($\eta_S$ and $\eta_L$) measured at 36C and 40.5C for healthy persons, and patients classed as WHO 4 and WHO 8 exhibiting no other underlying conditions (see Supplementary Table 2 and Methods). Standard errors are shown in parenthesis.

#### Supplementary Figures

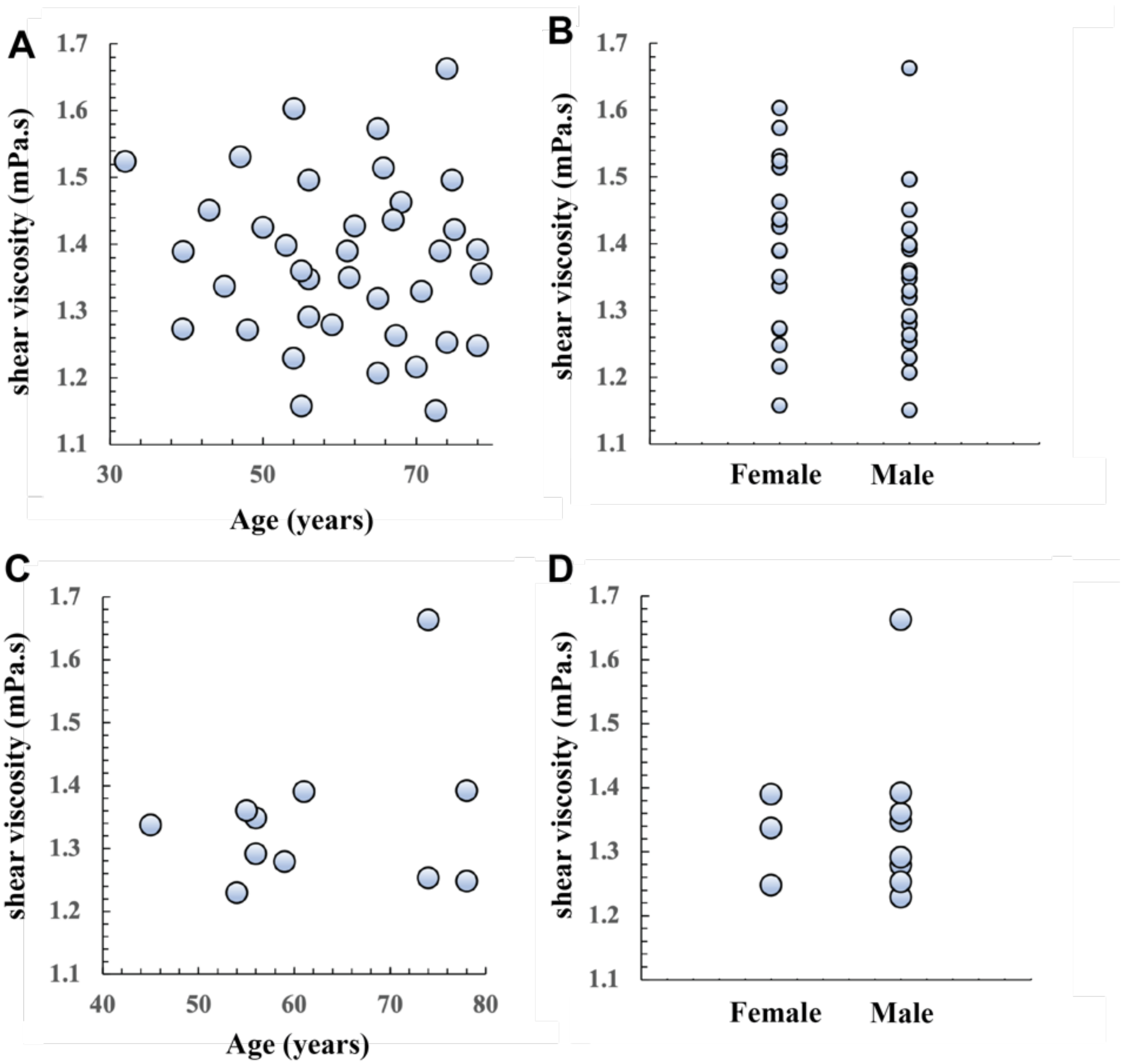


**Supplementary Fig 1: Age and gender dependence of viscosity. (A)** Age, and **(B)** gender dependence of the shear viscosity ($\eta_S$) of all obtained samples (Supplementary Table 2) measured at 36C, suggesting that there is no significant correlation of age or gender to the shear viscosity. **(C)** and **(D)** show age and gender dependence of the shear viscosity ($\eta_S$) of a subset of COVID patient samples used to perform temperature sweeps and longitudinal viscosity studies on (ones with no other identified underlying symptoms, see Main text and Supplementary Table 2). Here too there is no apparent correlation of age or gender to the viscosity.

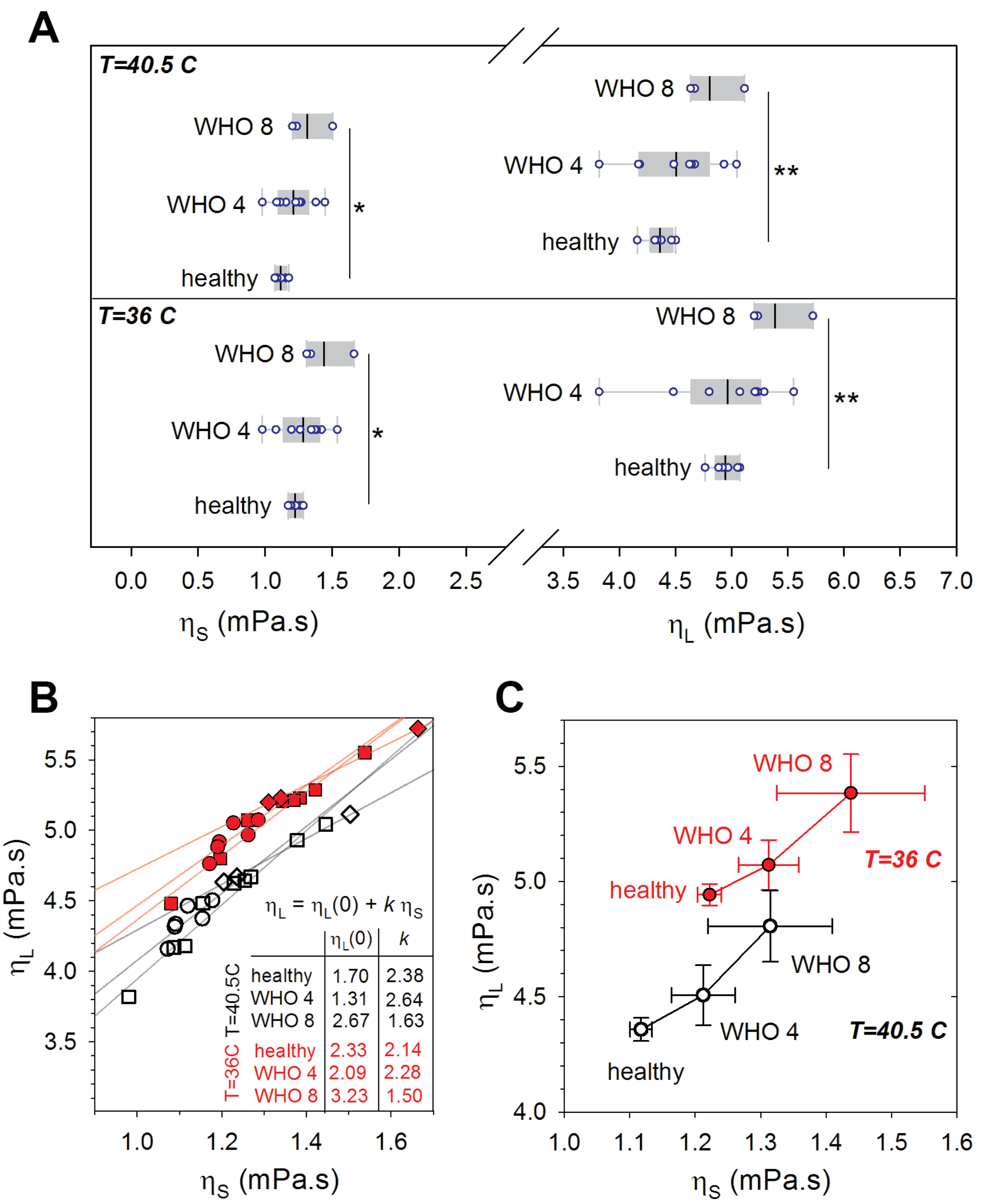


**Supplementary Fig 2: Differences and scaling of shear ($\eta_S$) and longitudinal ($\eta_L$) viscosity in healthy and COVID-patient plasma. (A)** $\eta_S$ and $\eta_L$ for the considered healthy and COVID-patient plasma (see Supplementary Table 2) at 36C and 40.5C. **(B)** Plot of $\eta_S$ and $\eta_L$ at 36C (red) and 40.5C (white) (*viz.* inset of Fig 2C), with linear fits for the different sample types. We note the slower increase of $\eta_L$ with respect to $\eta_S$ in WHO 8 patients ($k$), which is consistent with the observed difference in the scaling of $\eta_L$ with respect to $\eta_S$ of individual WHO 8 samples during temperature sweeps (Fig. 2C). **(C)** Mean values of $\eta_S$ versus $\eta_L$ for different indicated sample types at 36C and 40.5C (error bars are standard errors). “ * ” and “ ** ” indicate a significance at the level $p<0.01$ and $p<0.001$ respectively (unpaired t-test with Welch’s correction).

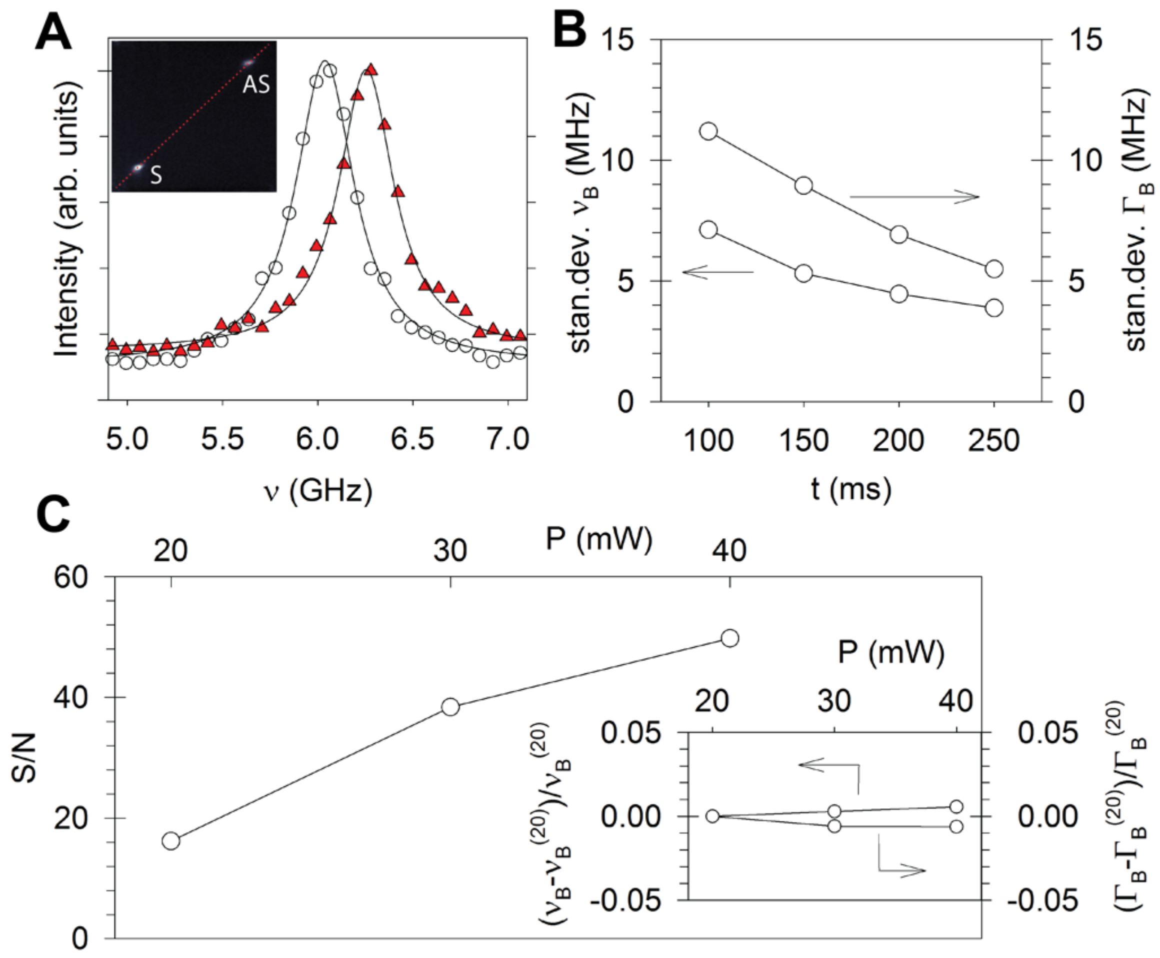


**Supplementary Fig 3: Precision and Signal-to-Noise ratio (S/N) of BLS measurements. (A)** Example of BLS Stokes peak measured at two temperatures fitted with a DHO (inset: raw spectral projection on detector array). (**B)** Precision of BLS frequency shift ($\nu_B$) and BLS linewidth ($\Gamma_B$) obtained from grid scans of water (as described in Methods) for different acquisition times per voxel (laser power at sample = 40mW). (**C)** S/N for different laser powers at the sample (exposure time 100ms) – see *Methods*. Inset shows relative change in the fitted $\nu_B$ and $\Gamma_B$ for the same range of laser powers, showing these are negligibly perturbed (<1%) and deviations due to laser sample heating can be considered insignificant.

### Supplementary Text:

#### Limitations:

***Refractive index and density:*** One limitation in extracting values of the longitudinal viscosity using BLS-spectroscopy is that it requires knowledge of the refractive index and mass density. By virtue of the Lorentz-Lorenz relation and derived Gladstone-Dale formula, the mass density can to a reasonable approximation be expressed in terms of the refractive index[1], which can be readily measured also as a function of temperature using commercial Abbe refractometers (as we have done here). This relies on the validity of the Lorentz-Lorenz relation and the values of the coefficients therein. While the former can reasonably be justified in dilute fluids such as plasma, regardless of precise protein composition[2], the validity of assuming the same proportionality coefficients between different samples can be questioned, since: (1) The contribution from lipids, which is known to vary significantly in plasma would have very different proportionality coefficients. Though typical lipid concentrations are on the order of ~1g/dL, in various diseases (such as untreated diabetes mellitus, lipoid nephrosis or hypothyroidism) concentrations can be orders of magnitude larger. For this reason we have in this study confined ourselves to patients who do not suffer other underlying conditions (including diabetes), however for practical diagnostic applications (where patients may have other underlying conditions) this needs to be considered. To this end complimentary chemical analysis or Raman spectroscopy measurements to identify lipid abundance could be considered. (2) The contribution from insoluble structures/suspensions on these coefficients is also unclear. It is certainly conceivable to employ Maxwell-Garnett (mixing) -like extensions to the Lorentz-Lorenz relation for suspension solutions, for this however further knowledge of their structural (optical, material, hydrodynamic) properties would be required.

These issues could be solved by directly also measuring the mass density. The problem is that direct measurement of mass density in small volumes, while possible using digital density meters employing e.g. the forced or pulsed oscillating U-tube principle[3] (e.g. used in Anton Paar, DMA 5000 M), still require >1mL in sample volume and an additional time consuming experimental step for each sample. To overcome this the potential of combining such a setup with a spectrometer (e.g. performing BLS directly on the plasma in the oscillating U-tube) is conceivable.

From a practical perspective (*i.e.* applications at point-of-care settings) the additional step of separately measuring the plasma refractive index using an Abbe refractometer is also undesirable. To this end one could obtain the refractive index by making use of the angle-dependence of BLS[4] or performing optical diffraction tomography,[5] that can both in principle be integrated with the BLS measurements in the same optical setup. Alternatively, given the reasonable transparency of plasma, one can also envision a custom design where the refractive index is calculated from precise measurements of changes in the refraction or attenuated total reflection angle of a second incident laser beam[6].

***Correctly obtaining the BLS linewidth:*** Due in part also to the relatively poor spectral resolution of BLS VIPA spectrometers compared to scanning multi-pass Fabry Perot spectrometers[7], care needs to be exercised to extract the true linewidth. In particular, deconvolution of the measured BLS spectra with the separately measured instrument spectral response should be carried out. It may be possible, to as a first approximation simply subtract the introduced broading (typically taken as the measured elastic scattering peak width) from the measured BLS linewidth[8]. While we have not done this here (and rather performed full spectral deconvolution), based on the almost constant amount that the linewidth is observed to decrease by deconvolution, this seems could also be a reasonable approach for correcting the linewidth. Regardless of the spectrometer employed it is also important to perform measurements at low numerical apertures to limit broading introduced by measuring different phonon wavevectors[9].

Finally, it may also be desirable to work with values normalized to those obtained for distilled water: namely defining a normalized longitudinal viscosity $\eta_L/\eta_L^w$, where $\eta_L^w$ is the longitudinal viscosity of distilled water measured under the same experimental conditions (in an analogous way to the normalized BLS frequency shift $\nu_B/\nu_B^w$ and linewidth $\Gamma_B/\Gamma_B^w$ introduced in[7]). The benefit of doing so is also that one would (to a first approximation) correct for systematic offsets due to instrument drift when operating under none-ideal conditions (but also other parameters used in the calculation of $\eta_L$), and thus obtain more robust values that correctly reflect variations/trends.

***Separating erythrocytes from plasma:*** The most time-consuming step between drawing patient blood and obtaining a measurement result in our workflow is the separation of erythrocytes from plasma (currently done by conventional laboratory centrifugation of blood collected in transport tubes, Methods). In practice (for patient monitoring) it may however be possible to completely skip this step, and one can envision periodic real-time monitoring of the blood viscosity of an individual in an ICU unit using a modified apheresis machine (akin to what is routinely used in plasma donations)[10]. This could be programed to every so often take small portions of blood, separate out the plasma (on which the BLS measurements are automatically performed), and return the remainder of the blood to the patient via intravenous tubes. Indeed, such measurements would allow one to measure at as near physiological conditions as possible and instantaneously after collection, avoiding any possible protein degradation that may occur over time and sample handling.

***Footprint of BLS setup:*** Our current BLS spectrometer and microscope setup is still rather bulky (~1m$^2$ on an optical bench). This is in order to allow us to work with commercially available optomechanical components and easily fine-tune the alignment. There is however no fundamental limitation why the spectrometer needs to be this large in a robust commercial device, and miniaturization of all optical components and folding of optical paths, to render an instrument a fraction of this size should be possible. Similarly, the use of a conventional microscopy stage is also not explicitly required, and one can readily envision the implementation of fiber coupled hand-held probes[11] (that can be used to measure directly in collection tubes), or install compact excitation/collection optics (in e.g. a micro-fluidic chamber) for real-time measurements in apheresis devices (see above).

***Supplementary References:***


1 Barer, R. Refractometry and Interferometry of Living Cells*†. *J. Opt. Soc. Am.* **47**, 545-556 (1957). https://doi.org:10.1364/JOSA.47.000545

2 Zhao, H., Brown, P. H. & Schuck, P. On the distribution of protein refractive index increments. *Biophys J* **100**, 2309-2317 (2011). https://doi.org:10.1016/j.bpj.2011.03.004

3 Oscillation-type density meters - Part 1: Laboratory instruments. **ISO 15212-1:1998**, www.iso.org/standard/28482.html (1998).

4 Fiore, A., Bevilacqua, C. & Scarcelli, G. Direct Three-Dimensional Measurement of Refractive Index via Dual Photon-Phonon Scattering. *Phys Rev Lett* **122**, 103901 (2019). https://doi.org:10.1103/PhysRevLett.122.103901

5 Schlüßler, R. *et al.* Correlative all-optical quantification of mass density and mechanics of subcellular compartments with fluorescence specificity. *eLife* **11**, e68490 (2022). https://doi.org:10.7554/eLife.68490

6 Born, M. & Wolf, E. *Principles of Optics*. 6 edn, (Pergamon Press, 1980).

7 Antonacci, G. *et al.* Recent progress and current opinions in Brillouin microscopy for life science applications. *Biophysical Reviews* (2020). https://doi.org:10.1007/s12551-020-00701-9

8 Yan, G., Bazir, A., Margueritat, J. & Dehoux, T. Evaluation of commercial virtually imaged phase array and Fabry-Pérot based Brillouin spectrometers for applications to biology. *Biomed. Opt. Express* **11**, 6933-6944 (2020). https://doi.org:10.1364/BOE.401087

9 Antonacci, G., Foreman, M. R., Paterson, C. & Török, P. Spectral broadening in Brillouin imaging. *Applied Physics Letters* **103**, 221105 (2013). https://doi.org:10.1063/1.4836477

10 Castillo, B., Dasgupta, A., Klein, K., Tint, H. & Wahed, A. in *Transfusion Medicine for Pathologists* (eds Brian Castillo *et al.*) 113-124 (Elsevier, 2018).

11 Pruidze, P., Chayleva, E., Weninger, W. J. & Elsayad, K. Brillouin scattering spectroscopy for studying human anatomy: Towards in situ mechanical characterization of soft tissue. *J. Eur. Opt. Society-Rapid Publ.* **19**, 31 (2023).
